# Lactation consultant support for breastfeeding people with HIV: HIV-knowledge, attitudes, stigma, and tele-lactation experiences in the United States and Canada

**DOI:** 10.1101/2025.08.05.25333056

**Authors:** Emily Barr, Lisa Abuogi, Mary Lingwall, Qian Qian, Leah Anthony, Joanna Vennekotter, Hulin Wu, Rebecca Tsusaki, Jennifer McKinney

## Abstract

**Background:** As infant feeding guidelines in the United States evolve to include shared decision-making approaches for pregnant and postpartum people with HIV (PP-PWH), lactation consultants (LCs) play an increasingly critical role in providing informed, compassionate support for breastfeeding/chestfeeding. However, limited knowledge and persistent stigma may hinder their preparedness and willingness to support PP-PWH in achieving their feeding goals.

**Methods:** We conducted a cross-sectional mixed-methods survey among 207 certified LCs in the United States and Canada. Quantitative data assessed HIV-related knowledge, stigma, and willingness to support breastfeeding in PP-PWH using adapted and validated instruments. Qualitative data from open-ended responses were thematically analyzed using the Health Stigma and Discrimination Framework.

**Results:** Participants demonstrated high general HIV knowledge (Mean = 9.29/10) but only moderate HIV breastfeeding knowledge (Mean = 10.8/21). Those with recent experience supporting PP-PWH in breastfeeding had significantly higher HIV breastfeeding knowledge (p < 0.001). Stigma levels were generally low, but stigma was significantly associated with more restrictive attitudes toward breastfeeding (p < 0.001). Qualitative findings revealed that LC willingness to support PP-PWH was shaped by perceived risks, personal comfort levels, professional ethics, understanding of transmission prevention, and systemic supports. Many emphasized respect for parental autonomy, nonjudgmental care, and the need for updated education and policy clarity.

**Conclusion:** LCs are motivated to support PP-PWH but face knowledge gaps and institutional barriers that must be addressed. Integrating targeted education, stigma reduction strategies, and peer-supported tele-lactation models may enhance LC confidence and improve equitable, person-centered care for families affected by HIV.

## Introduction

Infant feeding guidelines for pregnant and postpartum people with HIV (PP-PWH) have recently changed in the U.S., shifting from strict recommendations against breastfeeding/chestfeeding (BF/CF) to a shared decision-making model that supports BF/CF when clinically appropriate and desired. ^[1, 2]^ As this option becomes more accessible, specialized lactation support offers a promising strategy to safely increase BF/CF among PP-PWH and their children. Lactation consultants (LCs) play a key role in helping parents navigate the clinical and practical challenges of infant feeding, and their support is associated with increased confidence in BF/CF. ^[2, 3]^ LC involvement has shown particular value among families with complex or chronic conditions, including HIV. ^[3–5]^

Care providers and ancillary infant feeding support team members, like LCs, may not be aware of updated guidelines or lack the HIV-specific knowledge required to support PP-PWH. ^[2, 6, 7]^ Accurate, current training is essential, as common lactation issues may carry increased risk for PP-PWH. ^[2, 8]^ With targeted education, LCs can offer specialized care, including via telehealth, to support diverse, vulnerable lactating populations. ^[5, 9, 10]^

Although in-person LC care remains common, barriers like provider shortages, geographic isolation, and time constraints have prompted interest in tele-lactation; real-time lactation support delivered via secure telehealth platforms. This model has been shown to increase exclusive BF/CF rates, reduce early cessation, and is well received by users. ^[11, 12]^ Telelactation has been associated with increased exclusive breastfeeding rates, reduced early cessation, and high user satisfaction, as summarized in a systematic review of primary care-based telehealth models. ^[9]^ Tele-lactation expands high-quality services to underserved regions, particularly for clinically complex and marginalized populations. ^[13–16]^ For PPWH, it may offer additional support in addressing both medical and psychosocial barriers, including stigma and discrimination. ^[17, 18]^

The present study assesses LCs’ general HIV knowledge, HIV BF/CF-specific knowledge, attitudes toward supporting BF/CF among PP-PWH, and their tele-lactation experience.

## Methods

### Study Design

This descriptive cross-sectional survey study employed an embedded parallel mixed-method design, a framework that integrates quantitative and qualitative data collected concurrently to gain a more comprehensive understanding of the research topic. ^[19]^ The survey was administered via REDCap ^[20]^ from February 23, 2024, to May 3, 2024, following approval from the Committee for the Protection of Human Subjects at The University of Texas Health Science Center at Houston (UTHealth Houston). The institutional review board reviewed the study in compliance with the Department of Health and Human Services regulations for the protection of human subjects (45 CFR Part 46) and HIPAA requirements. A waiver of consent was granted for survey participation.

### Participants

Eligible participants were certified as lactation consultants or counselors for at least one year, had at least two years of experience supporting clients, and were fluent in English. Participants were 18 years or older, working in Canada or the United States and were recruited through multiple lactation support networks and professional platforms, including the International Lactation Consultant Association and Canadian Lactation Consultant Association websites, La Leche League, the African American Breastfeeding Network, state and provincial breastfeeding organizations, as well as LinkedIn and Instagram professional profiles. Participants were informed that the survey was anonymous, required approximately 20 minutes, and did not collect protected health information. Compensation was provided via an electronic $20 gift card.

### Survey Instruments

The survey assessed participants’ knowledge, attitudes, and experiences related to HIV, BF/CF, telehealth experience, and HIV-related stigma in maternal and infant care. It included both closed- and open-ended questions. Items were drawn from or adapted from validated tools, including: The HIV Knowledge Questionnaire, ^[21]^ the Telehealth Usability Questionnaire, ^[22]^ and the HIV Healthcare Provider Stigma Scale, ^[23]^ which was adapted for maternal-infant HIV care and lactation contexts. To assess participants’ knowledge of breastfeeding in the context of HIV, a team of clinical and research experts in lactation, pediatric and maternal HIV care, and public health developed a 10-item multiple-choice questionnaire with a maximum possible score of 21. The measure assessed core and specialized knowledge needed for lactation support in the context of HIV, addressing scenarios such as referral to HIV specialists, the role of maternal viral suppression, and when to pause breastfeeding for reassessment. It aimed to identify knowledge gaps and inform future training in evidence-based, person-centered care for families affected by HIV.

### Quantitative Analysis

Quantitative Data analysis was conducted using R (R version 4.0.4) and RStudio (version 2023.09.0+463). ^[24]^ Descriptive statistics were reported as means and standard deviations for continuous variables, and frequencies with percentages for categorical variables. The Kruskal-Wallis Test was used to assess differences in numeric variables across more than two groups, while the Wilcoxon rank-sum test was used to compare two groups. Cronbach’s Alpha was calculated to evaluate the internal consistency of three scales: HIV Knowledge scale, HIV Breastfeeding Knowledge scale, and HIV Stigma scale. To account for multiple comparisons, the False Discovery Rate (FDR) correction was applied using the Benjamini-Hochberg procedure.^[25]^

### Qualitative Analysis

We conducted a thematic analysis of free-text responses to explore participants’ attitudes and experiences related to supporting breastfeeding among PP-PWH. This method is well-suited for identifying patterns within qualitative data while allowing flexibility in interpretation. ^[26]^ The analysis was guided by the Health Stigma and Discrimination Framework, which considers stigma as a multi-level phenomenon, including individual, interpersonal, community, and structural dimensions, and its’ influence on health behaviors and care delivery. ^[27]^

Textual data were uploaded into MAXQDA for coding and theme development. A structured, team-based approach was used, informed by the FORT-CAST framework, which emphasizes collaborative coding and cross-checking for rigor and transparency. ^[28]^ Initial coding was conducted by one researcher and refined by a second. A third synthesized themes and identified cross-cutting patterns, while a fourth aligned final themes with the theoretical framework for coherence and depth. Themes were reviewed by clinicians and researchers, including a midwife, pediatric nurse practitioner, pediatrician, maternal-fetal medicine specialist, nurses, and lactation consultants, many with expertise in pediatric and maternal HIV. This validation process enhanced the credibility and transferability of findings, consistent with qualitative research best practices. ^[29]^

The Health Stigma and Discrimination Framework helped contextualize participants’ responses, illustrating how stigma-related drivers, facilitators, and institutional norms shape LC willingness to support PP-PWH. This structured process highlighted the complex, multilayered influences on provider attitudes and generated actionable insights to guide future training, policy, and systems-level interventions.

## Results

### Demographics

The sample of LCs (N=207) included in this study (Table 1) were predominately female (98.6%, n=204), non-Hispanic White (64.3%, n=133), Black (8.2%, n=17), Hispanic/Latino (5.3%, n=11), and were aged 36–50 years (48.3%; n=100). In the past two years, only 24.6% (n=51) had supported at least one PP-PWH. Experience with telehealth was common in our sample of LCs, with 95.7% (n=198) reporting previous experience with tele-lactation.

**Table 1.** Demographics, Race/Ethnicity, and Professional Characteristics of Lactation Consultant Participants (N = 207)

| **Characteristic** | **n (%)** |
| --- | --- |
| **Age** |  |
| 18–35 | 29 (14.01) |
| 36–50 | 100 (48.31) |
| 51 or older | 78 (37.68) |
| **Country of Practice** |  |
| United States | 180 (89.96) |
| Canada | 27 (13.04) |
| **Years of Experience as LC** |  |
| 1–5 years | 40 (19.32) |
| 6–10 years | 48 (23.19) |
| More than 10 years | 119 (57.5) |
| **Primary Practice Area** |  |
| Rural | 23 (11.11) |
| Suburban | 87 (42.03) |
| Urban | 43 (20.77) |
| All settings | 54 (26.09) |
| **Gender** |  |
| Female | 204 (98.55) |
| Male | 1 (0.48) |
| Non-binary | 1 (0.48) |
| Prefer not to answer | 1 (0.48) |
| **Race/Ethnicity** |  |
| White | 150 (72.46) |
| Black or African American | 17 (8.21) |
| Asian | 7 (3.38) |
| Hispanic or Latino/a | 11 (5.31) |
| Multiracial | 10 (4.83) |
| Other | 6 (2.89) |
| Prefer not to answer | 6 (2.89) |
| **Education Level** |  |
| Pre-Bachelor's | 29 (14.00) |
| Bachelor's Degree | 106 (51.20) |
| Post-Bachelor's (master's or higher) | 63 (30.43) |
| **Experience Supporting BF in PWH (past 2 years)** |  |
| No clients with HIV | 154 (74.39) |
| 1–5 clients with HIV | 51 (24.63) |
| More than 20 clients with HIV | 2 (0.96) |
| **The last time they heard about infant feeding guidelines in PP-PWH in North America** |  |
| Within the last year | 132 (63.73) |
| In the past 1–2 years | 17 (8.21) |
| Over 2 years ago or never heard | 58 (28.01) |
| **Telehealth Experience** |  |
| Yes | 198 (95.65) |
| No | 9 (4.34) |

**Note:** Participants could select more than one race/ethnicity. Notably, 133 (64.3%) participants identified as only White and non-Hispanic, highlighting the demographic homogeneity of the sample

### Score Summary

The HIV General Knowledge Scale (10 items, max score 10) ^[21]^ had a high mean score of 9.29 (SD = 0.94), indicating high overall knowledge, but demonstrated low internal consistency (Cronbach’s α = 0.42; average inter-item correlation = 0.07). Similarly, The HIV Stigma Scale (9 items, max score 36) ^[23]^ had a mean score of 26.2 (SD = 3.21) (higher score correlates to lower levels of HIV-related stigma) and moderate internal consistency (α = 0.65; inter-item correlation = 0.17). The HIV Breastfeeding Knowledge Scale (21 items, max score 21), developed by the study team specifically for this study, had a mean of 10.8 (SD = 4.11) and showed stronger internal consistency (α = 0.76; inter-item correlation = 0.13), indicating acceptable reliability for this new scale.

### Knowledge and Experience

Participants who had supported PP-PWH in the past two years (n = 53, 25.6%) had significantly higher HIV Breastfeeding Knowledge scores than those who had not (12.6 vs. 10.2, p < 0.001), but no significant differences were found in HIV General Knowledge scores or HIV-related stigma levels (Table 2). No significant differences in HIV Breastfeeding Knowledge, General HIV Knowledge, or HIV Stigma scores were observed based on years of LC experience (Table 2).

**Table 2.**
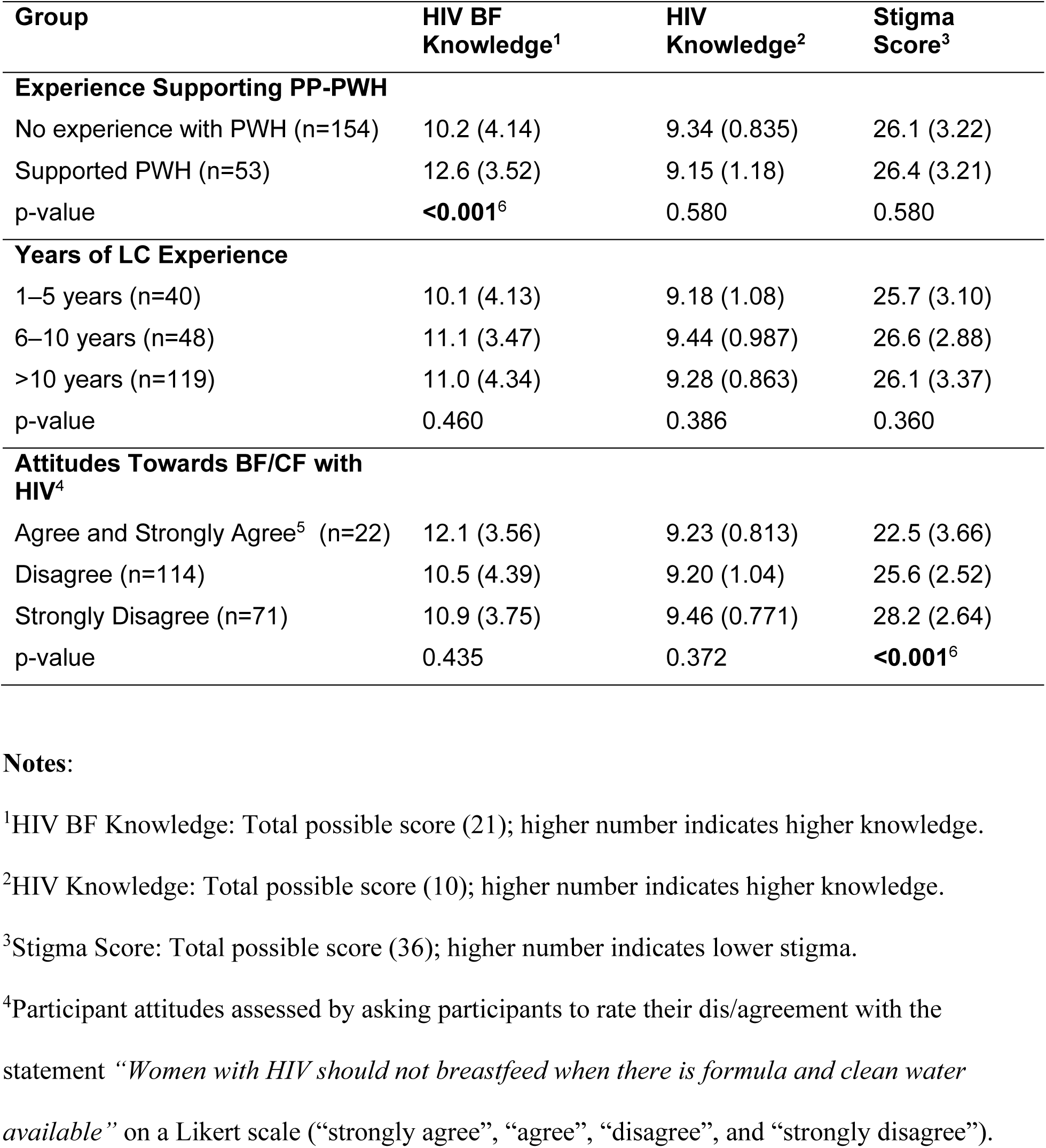
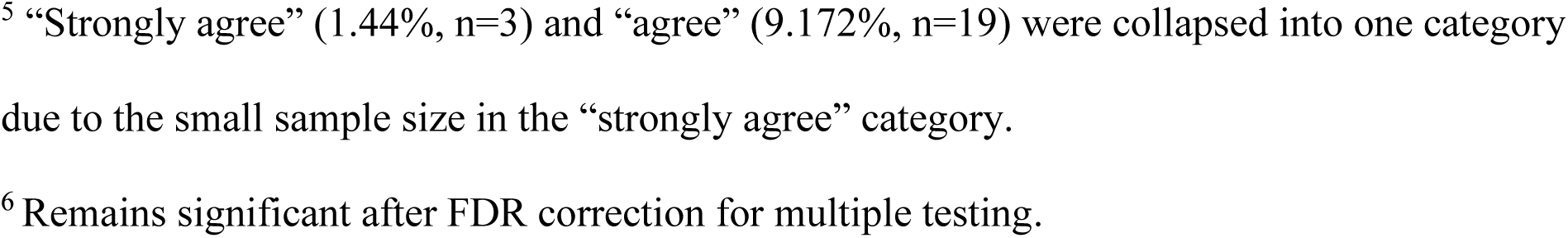
Differences in HIV Knowledge, HIV Breastfeeding Knowledge, and HIV Stigma Scores by Experience Supporting PWH, Years of Lactation Experience, and Attitudes Towards BF/CF with HIV.

| **Group** | **HIV BF Knowledge<sup>1</sup>** | **HIV Knowledge<sup>2</sup>** | **Stigma Score<sup>3</sup>** |
| --- | --- | --- | --- |
| **Experience Supporting PP-PWH** |  |  |  |
| No experience with PWH (n=154) | 10.2 (4.14) | 9.34 (0.835) | 26.1 (3.22) |
| Supported PWH (n=53) | 12.6 (3.52) | 9.15 (1.18) | 26.4 (3.21) |
| p-value | **<0.001<sup>6</sup>** | 0.580 | 0.580 |
| **Years of LC Experience** |  |  |  |
| 1–5 years (n=40) | 10.1 (4.13) | 9.18 (1.08) | 25.7 (3.10) |
| 6–10 years (n=48) | 11.1 (3.47) | 9.44 (0.987) | 26.6 (2.88) |
| >10 years (n=119) | 11.0 (4.34) | 9.28 (0.863) | 26.1 (3.37) |
| p-value | 0.460 | 0.386 | 0.360 |
| **Attitudes Towards BF/CF with HIV<sup>4</sup>** |  |  |  |
| Agree and Strongly Agree<sup>5</sup> (n=22) | 12.1 (3.56) | 9.23 (0.813) | 22.5 (3.66) |
| Disagree (n=114) | 10.5 (4.39) | 9.20 (1.04) | 25.6 (2.52) |
| Strongly Disagree (n=71) | 10.9 (3.75) | 9.46 (0.771) | 28.2 (2.64) |
| p-value | 0.435 | 0.372 | **<0.001<sup>6</sup>** |

**Notes:**
<sup>1</sup>HIV BF Knowledge: Total possible score (21); higher number indicates higher knowledge.
<sup>2</sup>HIV Knowledge: Total possible score (10); higher number indicates higher knowledge.
<sup>3</sup>Stigma Score: Total possible score (36); higher number indicates lower stigma.
<sup>4</sup>Participant attitudes assessed by asking participants to rate their dis/agreement with the statement “*Women with HIV should not breastfeed when there is formula and clean water available*” on a Likert scale (“strongly agree”, “agree”, “disagree”, and “strongly disagree”).
<sup>5</sup> “Strongly agree” (1.44%, n=3) and “agree” (9.172%, n=19) were collapsed into one category due to the small sample size in the “strongly agree” category.
<sup>6</sup> Remains significant after FDR correction for multiple testing.

### Attitudes Towards Autonomy and Stigma

Overall, LCs in our sample reported low levels of HIV-related stigma, with most expressing comfort with routine infant care and physical contact with people with HIV (Table 3). However, attitudes toward breastfeeding among PP-PWH varied and were significantly associated with stigma levels (Table 4). Overall, most of the cohort was supportive of autonomy for infant feeding choice and endorsed positive attitudes toward BF/CF with HIV (Table 2). However, many respondents felt that PP-PWH should disclose their status to sexual partners and/or the infant’s co-parent (Table 3).

**Table 3.** Attitudes and practices among lactation consultants towards people living with HIV (N = 207)

| **Attitude/Practice** | **n (%)** |
| --- | --- |
| Not worried about changing diapers of infant with HIV | 163 (78.7%) |
| Not worried about handling expressed breastmilk from person with HIV | 139 (67.1%) |
| Never avoided physical contact with patient due to HIV | 113 (54.6%) |
| Never wore double gloves | 95 (45.9%) |
| Never used special precautions (not used with other patients) | 84 (40.6%) |
| Never witnessed or engaged in neglect during delivery/postpartum | 73 (35.3%) |
| Belief: PWH should disclose to sexual partners | 202 (97.6%) |
| Belief: PWH should disclose to infant's co-parent | 178 (86.0%) |

**Note:.** Responses reflect the percentage and number of participants (out of 207) who reported either: (1) **non-stigmatizing practices** (e.g., answering “Never” or “Not at all” to stigma-related behavior questions), or (2) **agreement with statements** supporting disclosure (e.g., agreeing or strongly agreeing that people with HIV should disclose their status to sexual partners or co-parents). All items were coded such that higher percentages reflect less stigmatizing attitudes or behaviors.

**Table 4.** Comparison of HIV Knowledge and Stigma Scores among respondents based on LC support of autonomy in PP-PWH infant feeding decision.

|  | More Supportive of<br>Autonomy in<br>Choice<sup>1</sup> | Less Supportive of<br>Autonomy in Choice<sup>1</sup> | P-value |
| --- | --- | --- | --- |
| Sample | (n = 169) | (n = 38) |  |
| **HIV Breastfeeding Knowledge Score (Range: 0–21)<sup>2</sup>** |  |  |  |
| Mean (SD) | 11.1 (4.14) | 9.71 (3.83) | 0.108 |
| Median [Min, Max] | 12.0 [0, 18] | 10.0 [1, 16] |  |
| **General HIV Knowledge Score (Range: 0–10)<sup>3</sup>** |  |  |  |
| Mean (SD) | 9.30 (0.92) | 9.29 (1.01) | 0.954 |
| Median [Min, Max] | 10.0 [5, 10] | 10.0 [5, 10] |  |
| **HIV Stigma Score (Range: 9–36)<sup>4</sup>** |  |  |  |
| Mean (SD) | 26.4 (3.03) | 25.0 (3.76) | **0.020<sup>5</sup>** |
| Median [Min, Max] | 27.0 [15, 36] | 25.0 [16, 34] |  |

**Note:**
<sup>1</sup>LC support of autonomy in PP-PWH choice to BF/CF was assessed by asking participants to rate their dis/agreement with the statement *"Given that we cannot completely eliminate risk of HIV transmission via breastmilk, people with HIV should be able to choose to breastfeed"* on a scale from 1 ("strongly disagree") to 10 ("strongly agree"). Agreement is represented as a dichotomous variable of "More Supportive of Autonomy in Choice " (scores 7 – 10), and "Less Supportive of Autonomy in Choice," (scores 1 - 6).
<sup>2</sup>HIV BF Knowledge: Total possible score (21); higher number indicates higher knowledge.
<sup>3</sup>HIV Knowledge: Total possible score (10); higher number indicates higher knowledge.
<sup>4</sup>Stigma Score: Total possible score (36); higher number indicates lower stigma.
<sup>5</sup>FDR correction was applied across 12 comparisons; results with $q < 0.05$ are considered significant.

**Table 5.** Selected questions from stigma and attitudes towards people with HIV scale compared to experience supporting people with HIV.

| **Question** | **No experience**<br>**supporting people**<br>**with HIV**<br>**(n=154)** | **Has some experience**<br>**supporting people**<br>**with HIV**<br>**(n=53)** |
| --- | --- | --- |
| **How worried are you about acquiring HIV from changing diapers of an infant exposed to HIV?** |  |  |
| Very worried | 1 (0.6%) | 0 (0%) |
| Somewhat worried | 10 (6.5%) | 1 (1.9%) |
| A little worried | 22 (14.3%) | 10 (18.9%) |
| Not worried at all | 121 (78.6%) | 42 (79.2%) |
| **How worried are you about acquiring HIV from handling breast milk from a patient living with HIV** |  |  |
| Very worried | 3 (1.9%) | 0 (0%) |
| Somewhat worried | 17 (11.0%) | 2 (3.8%) |
| A little worried | 33 (21.4%) | 13 (24.5%) |
| Not worried at all | 101 (65.6%) | 38 (71.7%) |
| **Women with HIV should not breastfeed when there is formula and clean water available** |  |  |
| Strongly agree | 0 (0%) | 3 (5.7%) |
| Agree | 16 (10.4%) | 3 (5.7%) |
| Disagree | 87 (56.5%) | 27 (50.9%) |
| Strongly disagree | 51 (33.1%) | 20 (37.7%) |
| **A person with HIV should disclose their HIV status to their sexual partners** |  |  |
| Strongly agree | 114 (74.0%) | 41 (77.4%) |
| Agree | 36 (23.4%) | 11 (20.8%) |
| Disagree | 0 (0%) | 0 (0%) |

| Question | No experience<br>supporting people<br>with HIV<br>(n=154) | Has some experience<br>supporting people<br>with HIV<br>(n=53) |
| --- | --- | --- |
| Strongly disagree | 4 (2.6%) | 1 (1.9%) |
| **A pregnant or postpartum**<br>**woman/person with HIV should**<br>**disclose their HIV status to the father**<br>**or other parent of their infant** |  |  |
| Strongly agree | 64 (41.6%) | 24 (45.3%) |
| Agree | 66 (42.9%) | 24 (45.3%) |
| Disagree | 21 (13.6%) | 3 (5.7%) |
| Strongly disagree | 3 (1.9%) | 2 (3.8%) |

**Note:** This table presents item-level comparisons of selected stigma and attitude questions
between lactation consultants with no experience supporting people with HIV in the previous 2 years.

Most participants strongly agreed with the statement "Given that we cannot completely eliminate risk of HIV transmission via breastmilk, people with HIV should be able to choose to breastfeed” (71.49% n=122) (Table 4). This item was used to create two dichotomous groups of those “More supportive of autonomy in choice” (scores 7-10 on Likert scale with 10 indicating strong agreement), and “less supportive of autonomy in choice” (scores 1-6). Notably only 5.8% responded with scores of 1-3, indicating an overall positive attitude towards autonomy in infant feeding decisions in this cohort. Participants who were more supportive of PP-PWH’s right to choose their feeding method were found to have significantly lower levels of HIV stigma (p < 0.020) and trended towards increased HIV Knowledge scores (p = 0.108). In contrast, there was no significant difference in General HIV Knowledge scores between the two groups (mean = 9.30 vs. 9.29; *p* = 0.954).

Attitudes toward BF/CF in the context of HIV were assessed using agreement with the statement, “Women with HIV should not breastfeed when there is formula and clean water available.” Most participants disagreed or strongly disagreed, reflecting generally permissive views toward BF/CF among PP-PWH. These attitudes were significantly associated with stigma levels but not with HIV knowledge (Table 2).

### Willingness to Learn

All but two LCs in the sample reported a willingness to learn more about HIV and BF/CF to provide lactation support to PWH (99%, n=203). Most participants expressed interest in providing tele-lactation services to PP-PWH following appropriate education (89.8%, n=185). The majority (92.8%, n=192) preferred continuing education credit, and 62.8% indicated interest in certification (n=130) as motivation to improve HIV-specific knowledge.

### Qualitative Results

LCs provided free-text responses on their willingness to support BF/CF among PP-PWH, reflecting on HIV transmission risk, BF/CF benefits, clinical experience, ethics, access to evidence-based knowledge, and institutional support. Many emphasized patient autonomy and the need for unbiased, informed care, calling for improved education and guidance over restrictive policies. Several highlighted the importance of expanding lactation support, including tele-lactation, to ensure equitable care for PP-PWH.

### LC Attitudes Toward BF/CF Among PP-PWH

Most LCs in our study reported that PP-PWH interested in BF/CF under appropriate medical care should be supported in their decision to BF/CF.

“We should encourage her to get appropriate treatment and make an educated choice, then to support her with her decision.” (age 36-50, 6-10 years’ experience, USA)

“This should be the choice of the family when presented the full information along with the guidance/consistent follow up of a care team for both parent and child.”

(age 36-50, >10 years’ experience, USA)

Other LCs described personal discomfort with supporting breastfeeding in high-resource settings where formula is available; while others reported feeling comfortable supporting PP- PWH with BF/CF, but only in specific maternal clinical contexts, such as an undetectable viral load.

“I wouldn’t feel comfortable telling the mother to breastfeed when we have safe water and formula options.” (age 36–50, >10 years’ experience, USA)

“If mother’s titers are undetectable and infant and mother are both compliant with retroviral medications, the family’s wish to breastfeed should be supported if consistent, optimal management can be provided.” (age >51, >10 years’ experience, USA)

“I would be ok to breastfeed if the virus is undetectable and the mother is in a stable place both physically and emotionally.” (age >51, 3-5 years’ experience, USA)

### Risks and Benefits

Many LCs in this sample repeated the view that infant feeding decisions should be based on a careful evaluation of the relative risks and benefits of available options and emphasized the ultimate authority of decision-making to the mother/parent/family.

“Honest and open sharing of the pros and cons so the mother can make up her own mind.” (age >51, >10 years’ experience, USA)

“It is our job to educate mothers about the risks specific to breastmilk transmission. We should encourage her to get appropriate treatment and make an educated choice, then to support her decision.” (age 36-50, 6-10 years’ experience, USA)

“Provide as much information as is available about risks and benefits so that they can make informed choices”. (age 36-50, 6-10 years’ experience, USA)

“I believe it is shared decision making. It is the HCP’s [healthcare provider’s] job to provide accurate information about the risk and benefits and the parents role to decide what is best for their family.” (age >51, >10 years’ experience, USA)

Though many LCs focused their risk assessment on the inherent risks of HIV transmission, some also stressed the importance of human breastmilk for infants.

“Risk of HIV transmission must be carefully weighed against the significant benefits of human milk.” (age 36-50, 6–10 years’ experience, USA)

“Perhaps the short and long-term benefits of breastfeeding to both infant and maternal health outweighs the very small risk. However, I also believe that parents have the right to make an informed decision and if they feel the risk is too great, we should support them in their decisions.” (age > 51, >10 years’ experience, USA)

### HIV Stigma & Unbiased Care

LCs in our sample shared a commitment to approaching BF/CF among PP-PWH without judgment, and shared concerns about the impact of HIV stigma on effective, equitable lactation care for PP-PWH who BF/CF. Respect for client autonomy, even when LCs report that they personally disagree, emerged as a theme throughout the free-text responses.

“This is entirely up to the parent. My opinion doesn’t matter. They need to be provided the risks of doing so (the risks of formula, the risks of not breastfeeding, the risk of HIV transmission) and weigh this against the benefits (for both of them), their personal feelings around feeding their baby and make an informed decision. How I feel is irrelevant.” (age 18-35, 3-5 years’ experience, Canada)

### Access to Care & Telehealth

LCs described telehealth as a valuable tool for improving equitable lactation care for PP- PWH, highlighting its ability to provide specialized support, reduce stigma through private at-home access, and increase reach for those in remote areas or without transportation.

“A person with HIV may be able to access a professional who specializes in HIV and provide a higher level of care to help the family meet their goals.” (age 18-35, 3-5 years’ experience, Canada)

“A patient may feel like they have more privacy. They can get care easily and confidentially.” (age >51, >10 years’ experience, USA)

“It is a great option if they live remote and I can’t drive to their home to support them.” (age >51, 1-2 years’ experience, USA)

“Potential for more affordable care accessible by people with transportation difficulties or distant from health care.” (age >51, 6-10 years’ experience, USA)

### Willingness to Learn

Several LCs expressed the desire for deeper knowledge on HIV and BF/CF and shared an interest in more guidance & protocols, trainings, and educational materials to provide on-going support to build HIV competencies in the LC workforce.

“I would like more education about HIV and breastfeeding… My training has never outlined techniques to safely breastfeed with HIV.” (age 36-50, 6–10 years’ experience, USA)

This analysis reveals that while many LCs are ready to support PP-PWH in breastfeeding, their confidence is shaped by knowledge, institutional clarity, and emotional readiness. Enhanced training, consistent guidelines, and culturally responsive peer support could increase willingness and improve outcomes for families affected by HIV.

## Discussion

This study identifies gaps and opportunities in LCs knowledge, attitudes, and preparedness to support BF/CF among pregnant and PP-PWH in North America, with attention to the role of tele-lactation in expanding access. While general HIV knowledge was high, HIV-specific breastfeeding knowledge was significantly lower and more robust among LCs with direct experience supporting PP-PWH. This aligns with prior research linking clinical exposure to more supportive attitudes and reduced stigma in HIV care. ^[30, 31]^

Encouragingly, nearly all participants expressed interest in learning more about HIV and lactation, with over 90% reporting willingness to provide tele-lactation support to PP-PWH following appropriate training. This presents a promising opportunity to develop accessible, targeted educational initiatives, particularly those offering continuing education credit or certification. Success in other lactation specialties, such as maternal diabetes and NICU care, suggests such training is both feasible and effective. ^[4]^

These patterns were echoed in the qualitative responses, where many LCs expressed a desire to support BF/CF while acknowledging uncertainty about clinical protocols, risks, and their own emotional readiness. Applying the Health Stigma and Discrimination Framework, ^[27]^ participant narratives revealed how stigma manifests through institutional ambiguity, provider discomfort, and inconsistent guidance. LCs with strong HIV-specific breastfeeding knowledge felt more confident, while those with less exposure relied on outdated policies or personal bias. Structural gaps in education and collaboration further limited affirming care.

Quantitative findings demonstrated that LCs with higher HIV breastfeeding knowledge had significantly lower stigma scores and were more likely to support PP-PWH’s autonomy in feeding choice. This association was mirrored in the qualitative data, where participants emphasized shared decision-making and ethical imperatives to center patient autonomy, even when personal beliefs conflicted with clinical evidence. Notably, many LCs described their role not as directing care but as empowering parents with accurate, nonjudgmental information to make informed choices.

Together, these findings illustrate the power of HIV-specific knowledge to mitigate stigma and shift provider attitudes toward relational, person-centered care. Nearly all LCs reported a willingness to engage in additional training, and many called for standardized protocols and interdisciplinary collaboration to better serve PP-PWH. Addressing these needs through structured, stigma-informed education and practice guidelines can support a more equitable care environment.

Tele-lactation emerged as a promising avenue for reducing barriers and stigma in accessing lactation care. LCs noted that virtual visits can offer privacy, confidentiality, and logistical convenience, particularly for those in rural or underserved. ^[32]^ Prior studies have documented the value of peer counselors and telehealth in improving outcomes for marginalized groups. ^[18]^ The HIV Breastfeeding Knowledge tool developed for this study demonstrated acceptable internal consistency and may serve as a useful metric in evaluating provider readiness to support PP-PWH. Integrating this tool into future training and evaluation efforts may strengthen workforce preparedness.

Applying the Health Stigma and Discrimination Framework helped contextualize how knowledge, experience, and institutional structures interact to shape LCs’ support for PP-PWH. These insights underscore the importance of combining stigma-reduction strategies, tailored education, and innovative care delivery models like telehealth to foster affirming, informed care for families affected by HIV.

Implications for research and clinical care include the need for continued evaluation of HIV-specific lactation education and its’ impact on both provider attitudes and patient outcomes. Findings underscore the need to integrate HIV-related lactation content into foundational education for lactation consultants, midwives, nurses, and perinatal providers. Training should emphasize stigma reduction, evidence-based care, and relational decision-making infant feeding.

Future studies should assess the effectiveness of training models that integrate relational decision-making and culturally responsive approaches. Clinically, structured peer support programs may be a valuable complement to LC services. Given the persistent stigma surrounding HIV, peer support models, particularly those delivered via telehealth, could provide culturally concordant, empathetic support that enhances trust and engagement. ^[2, 8]^ Such models have demonstrated success in breastfeeding support and may hold promise for improving lactation support for PP-PWH. ^[33, 34]^

## Limitations

This cross-sectional, self-reported study limits causal inference and may introduce social desirability bias, especially in responses about stigma and clinical behaviors. The sample was mostly U.S.-based, limiting generalizability across North America. The general HIV knowledge scale, while validated, is older and not tailored to lactation contexts. In contrast, the HIV Breastfeeding Knowledge scale was developed for this study and showed strong internal consistency but should be further tested in diverse samples. The qualitative data came from free-text responses and in-depth interviews may have provided more detail on the experiences and attitudes of the LCs in the field.

## Conclusion

LCs are motivated to better support PP-PWH but require specialized knowledge, institutional support, and greater confidence to do so effectively. To advance equitable lactational care, thoughtfully designed educational programs, clear and inclusive national guidelines, and innovative care models are essential. Expanding access to lactation care through tele-lactation and other remote support options is also critical, particularly for reaching underserved families particularly in rural or highly congested urban areas. Collectively, these strategies can ensure equitable, person-centered lactation support across the perinatal continuum, regardless of HIV status.

## Data Availability

All data produced in the present study are available upon reasonable request to the authors

## Author Approvals

I have the right to post this manuscript and confirm that all authors have assented to posting of the manuscript and inclusion as authors.

## Competing Interests & Financial Disclosure

The authors have no competing interests to disclose or financial disclosures to declare.

## Funding Statement: Funding Information

This work was supported by the Sumner Roy Kates Charitable Trust and the Cizik School of Nursing at UTHealth Houston Dean’s Award.

## Notes

### Competing Interest Statement

The authors have declared no competing interest.

### Author Declarations

The survey was administered via REDCap from February 23, 2024, to May 3, 2024, following approval from the Committee for the Protection of Human Subjects at The University of Texas Health Science Center at Houston (UTHealth Houston). The institutional review board reviewed the study in compliance with the Department of Health and Human Services regulations for the protection of human subjects (45 CFR Part 46) and HIPAA requirements. A waiver of consent was granted for survey participation.

